# Improvements in the incidence and survival of cancer and cardiovascular but not infectious disease have driven recent mortality improvements in Scotland: nationwide cohort study of linked hospital admission and death records 2001–2016

**DOI:** 10.1101/19003012

**Authors:** Paul RHJ Timmers, Joannes J Kerssens, Jon W Minton, Ian Grant, James F Wilson, Harry Campbell, Colin M Fischbacher, Peter K Joshi

**Affiliations:** Centre for Global Health Research, Usher Institute of Population Health Sciences and Informatics, University of Edinburgh, Edinburgh, UK; Information Services Division, NHS National Services Scotland, Edinburgh, UK; Public Health Observatory, NHS Health Scotland, Edinburgh, UK; MRC Human Genetics Unit, Institute of Genetics and Molecular Medicine, University of Edinburgh, Edinburgh, UK

## Abstract

**Objectives:** To identify the causes and future trends underpinning improvements in life expectancy in Scotland and quantify the relative contributions of disease incidence and survival.

**Design:** Population-based study.

**Setting:** Linked secondary care and mortality records across Scotland.

**Participants:** 1,967,130 individuals born between 1905 and 1965, and resident in Scotland throughout 2001–2016.

**Main outcome measures:** Hospital admission rates and survival in the five years following admission for 28 diseases, stratified by sex and socioeconomic status.

**Results:** The five hospital admission diagnoses associated with the greatest burden of death subsequent to admission were “Influenza and pneumonia”, “Symptoms and signs involving the circulatory and respiratory systems”, “Malignant neoplasm of respiratory and intrathoracic organs”, “Symptoms and signs involving the digestive system and abdomen”, and “General symptoms and signs”. Using disease trends, we modelled a mean mortality hazard ratio of 0.737 (95% CI 0.730–0.745) across decades of birth, equivalent to a life extension of ∼3 years per decade. This improvement was 61% (30%–93%) accounted for by improvements in disease survival after hospitalisation (principally cancer) with the remainder accounted for by a fall in hospitalisation incidence (principally heart disease and cancer). In contrast, deteriorations in the incidence and survival of infectious diseases reduced mortality improvements by 9% (∼3.3 months per decade). Overall, health-driven mortality improvements were slightly greater for men than women (due to greater falls in disease incidence), and generally similar across socioeconomic deciles. We project mortality improvements will continue over the next decade but will slow down by 21% because much of the progress in disease survival has already been achieved.

**Conclusion:** Morbidity improvements broadly explain observed improvements in overall mortality, with progress on the prevention and treatment of heart disease and cancer making the most significant contributions. The gaps between men and women’s morbidity and mortality are closing, but the gap between socioeconomic groups is not. A slowing trend in improvements in morbidity may explain the stalling in improvements of period life expectancies observed in recent studies in the UK. However, our modelled slowing of improvements could be offset if we achieve even faster improvements in the major diseases contributing to the burden of death, or if we improve prevention and survival of diseases which have deteriorated recently, such as infectious disease, in the future.

**Summary box:** *What is already known on this topic:*

- Long term improvements in Scottish mortality have slowed down recently, while life expectancy inequalities between socioeconomic classes are increasing.
- Deaths attributed to ischaemic heart disease and stroke in Scotland have declined in the last two decades.

*What this study adds:*

- Gains in life expectancy can largely be attributed to improvements in cancer survival and falls in incidence of cancer and cardiovascular disease.
- The hospitalisation rate and survival of several infectious diseases have deteriorated, and for urinary infections, this decline has been more rapid in more socioeconomically deprived classes.
- Improvements in morbidity are projected to slow down, with much progress in survival of heart disease and cancer already achieved, and align with the recently observed slow-down in mortality improvements.

## Introduction

In recent decades, there has been a substantial improvement in life expectancies at birth in the UK(1). More recently, several studies have suggested that there has been slowdown in improvements in the USA, UK, France, Germany, Sweden, the Netherlands and other Organisation for Economic Co-operation and Development countries; however, the causes are less clear, with speculation that they may arise from slowing improvements in cardiovascular disease, increased influenza mortality and/or pressure on health and social care services(1–8). Understanding trends in disease incidence and subsequent survival could illuminate such trends in mortality, and disentangling how and how much different diseases contribute has the potential to reveal whether investment in healthcare and research is directed at the most urgent diseases and most affected individuals.

Through its electronic Data Research and Innovation Service (eDRIS), Scotland has linkage of historical individual death and electronic health records in a controlled environment, with specific study approvals by the Public Benefit and Privacy Panel. This allows direct modelling at an individual level of the incidence of disease and subsequent death or survival of subjects. Furthermore, because historic records are available and the whole population is covered, a retrospective cohort study can be constructed (with inherent representativeness of the initial sample, with very complete levels of follow-up, and without survivor bias).

Here, we use population-wide data between 2001 and 2016 on residents of Scotland born before 1966 to explore how trends in longevity were driven by different trends in broad classes of disease incidence or survival, and highlight diseases which have shown more or less improvement in their contribution to overall mortality. We partition overall mortality by sex and socioeconomic status and, assuming past disease improvements continue to the same extent in the future, use these results to project future improvements in mortality and their changing sources.

## Methods

### Data sources

We received administration and care records from NHS National Services Scotland (NSS) from 2001. All methods and results are reported in line with RECORD guidelines(9).

#### Community Health Index dataset

Records were extracted from the historic and current Community Health Index (CHI) dataset. This is a register of all patients in NHS Scotland and is fed by eight regional databases (e.g, GP database, cancer screening). The register is considered complete from 2001 onwards. The CHI number, contained in the dataset, is effectively a patient identifier and added to other health datasets to make linkage possible, for instance between hospital admissions, death records and the Scottish cancer registry(10). Our extract consisted of 2,691,304 de-identified records, constituting the identified population of Scotland in 2001 who had been born between 1905 and 1965. The Scottish Index of Multiple Deprivation (SIMD)(11) was used to quantify socioeconomic status, determined by individuals’ full postcode, and subsequently converted into deciles. The dataset we received contained only records with district-level postcodes and SIMD deciles, of which we excluded individuals with district codes with less than 5,000 individuals (thereby excluding anomalous postcodes, often with special meanings, such as “marketing campaign”; N = 11,564). We also excluded individuals missing from the CHI database in 2016, but not recorded as dead (and therefore likely transferred out of Scotland; N = 573,711), individuals with record discrepancies between the CHI and National Records of Scotland databases (N = 79,131), and individuals with records outside of the study dates or missing information on socioeconomic class (N = 59,767), giving 1,967,130 individuals for analysis after quality control (S Figure 1). Characteristics of the excluded individuals were similar to the rest of the population, except for postcode exclusions and database transfers, which were missing socioeconomic information and death records, respectively, as expected (S Figure 1—Source Data 1).

**Figure 1a:**
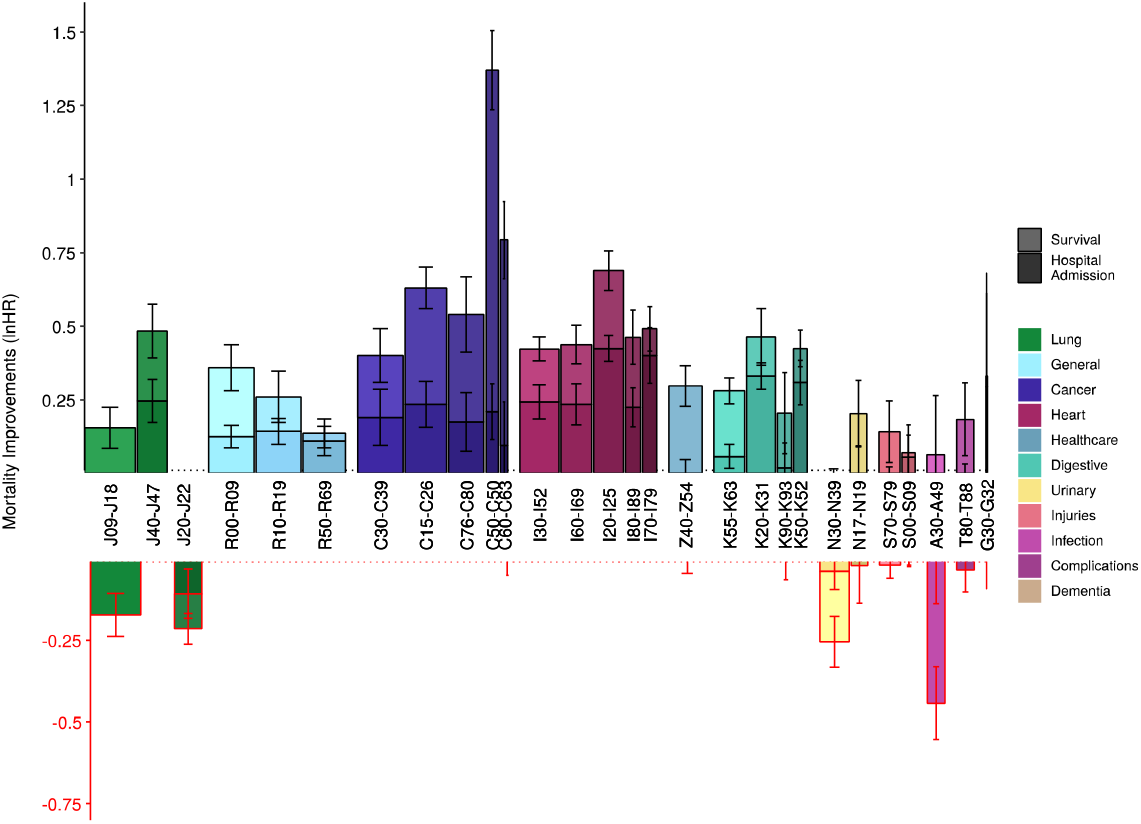
Modelled decade-of-birth upon previous decade-of-birth hospitalisations and survival show large improvements in cancer survival and heart disease incidence but deteriorations in infectious disease. Bars represent the mean improvements in hospital admission rate across decades of birth (darker bars), added to changes from 2001 to 2011 in 5-year survival rates following hospital admission (lighter bars). Both measures are expressed in age-adjusted lnHR. For definitions of each ICD10 block, see Table 2. Width of the bars represents the relative burden of death of each disease based on total first-time hospital admissions and 5-year mortality; as such, the total area of each bar represents the relative contribution to improvements – or deteriorations – in population mortality. Error bars are standard errors of the Cox model coefficient. G30–G32 had too few hospital admissions to accurately model improvements (Survival: lnHR 0.04, SE 0.14; Hospital admission lnHR 0.29, SE 0.20). Z40-Z54 only showed improvements in survival.

**Figure 1b:**
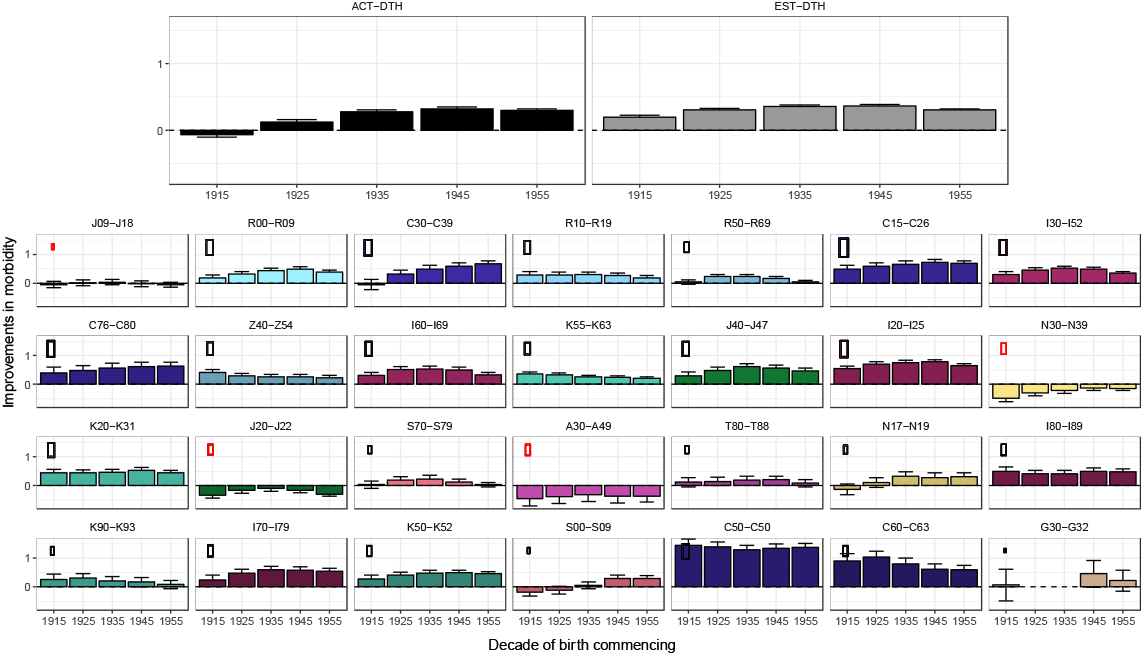
Modelled decade-of-birth upon previous decade-of-birth mortality reductions due to morbidity changes broadly track observed trends in mortality. Panels represent the combined improvements in hospital admission rate for each ICD10 disease block (for definitions see Table 2) and 5-year mortality rates following hospital admission, expressed in age-adjusted lnHR, split by decade of birth - under the model where change in incidence of disease is modelled by decade of birth and added to the survival effect is the change in subsequent 5 year survival rates from incidences in 2001 and 2011. Dots represent the relative contribution of the disease to the overall improvements in morbidity driven mortality. This is summarised by EST-DTH: modelled trend in deaths based on morbidity-grey bars and calculated as the weighted effect of the coloured bars), with larger dots indicating a greater contribution to morbidity improvements. A red circle around the dot indicates a negative contribution (i.e. deterioration). The black bars (ACT-DTH: observed trend in actual deaths) represent the improvements in mortality observed from death records, by decade of birth, for comparison. Diseases have been ordered by their burden of death (Table 2), so smaller bars in early panels may have similar effect on the grey bar average (indicated by the dot size) as larger bars in later panels. See Supplementary Data File 2 for this graph stratified by sex and deprivation.

#### NRS death records

We received 1,477,796 death records from the National Registry of Scotland, of which 699,093 could be matched to the CHI database before quality control. Of the matched records, 176,197 belonged to individuals that were excluded during CHI quality control, leaving 602,506 total deaths for analysis (Table 1).

**Table 1:** Description of the data. The population included almost 2 million individuals (one-third of whom died during the study). Over 90% of deceased individuals had a hospital admission record. Median projected age at death (under Kaplan-Meier curves) for males is 71.1 and 82.2, in the most and least deprived areas, respectively (females 76.6 and 85.2). See Table 1—Source Data 1 for descriptives by deprivation including ICD10 codes. N – Number of records; Admitted – Individuals admitted to hospital at least once; Hospital Visits – All records of visiting the hospital; Age Entry – Age at the start of the study period (1 December 2000); Age Exit – Age at the end of the study period (31 January 2016) or at the end of life.

| Sex | Dead | N |  |  | Age Entry |  |  | Age Exit |  |  | Hospital Visits |  |  |
| --- | --- | --- | --- | --- | --- | --- | --- | --- | --- | --- | --- | --- | --- |
|  |  | Individuals | Admitted | Hospital Visits | Mean | Median | SD | Mean | Median | SD | Mean | Median | SD |
| Male | FALSE | 633,953 | 429,659 | 2,315,915 | 50.9 | 49.4 | 10.3 | 65.6 | 64.3 | 9.8 | 3.7 | 2.0 | 6.8 |
| Male | TRUE | 283,835 | 283,835 | 2,742,686 | 68.0 | 69.3 | 11.7 | 75.6 | 77.1 | 11.4 | 9.7 | 7.0 | 11.4 |
| Female | FALSE | 730,671 | 511,632 | 2,764,040 | 52.7 | 51.1 | 11.6 | 67.4 | 66.1 | 10.8 | 3.8 | 2.0 | 7.0 |
| Female | TRUE | 318,671 | 318,671 | 2,895,443 | 72.1 | 73.8 | 12.0 | 79.7 | 81.7 | 11.5 | 9.1 | 6.0 | 11.0 |
| Both | FALSE | 1,364,624 | 941,291 | 5,079,955 | 51.9 | 50.3 | 11.1 | 66.6 | 65.3 | 10.4 | 3.7 | 2.0 | 6.9 |
| Both | TRUE | 602,506 | 602,506 | 5,638,129 | 70.1 | 71.7 | 12.0 | 77.8 | 79.5 | 11.6 | 9.4 | 6.0 | 11.2 |
| Both | ALL | 1,967,130 | 1,543,797 | 10,718,084 | 57.5 | 55.4 | 14.1 | 70.0 | 69.2 | 11.9 | 5.4 | 3.0 | 8.8 |

**Table 2:**
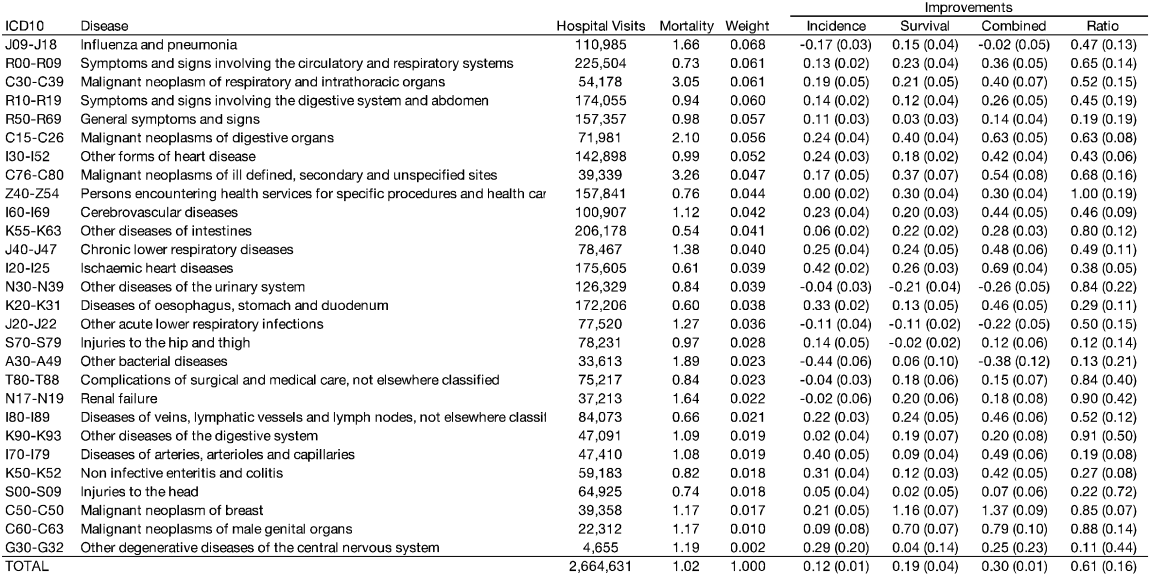
Relative mortality burden of hospital admission by disease grouping and improvements in hospitalisation incidence and survival. ICD10 – Diseases contained within the disease grouping, coded by International Classification of Disease Codes, Tenth Revision. See Table 1—Source Data 2 for counts of 3-letter ICD10 records within each ICD10 block. Disease – Description of the diseases within the disease grouping. Hospital visits – Number of first-time admissions with main diagnosis falling within the disease block. Mortality – Mortality compared to individuals who had not yet or ever been admitted for the disease group (in lnHR). Weight – Relative burden of death, calculated as hospital admissions multiplied by the mortality in the first five years after admission and scaled to [0-1]. Incidence – Improvements in disease incidence as captured by mean differences in hospital admissions between decades of birth (lnHR), with negative numbers indicating a deterioration. Survival – Improvements in 5-year mortality hazard following admission between 2001– 2011 (lnHR). Combined – linear combination of improvements in disease incidence and survival. Ratio – the ratio of improvements in disease survival to incidence of hospital admission. Standard errors are listed in parentheses. See Table 2—Source Data 1 for these data by sex and deprivation. See Table 2—Source Data 2 for the relative burdens of all disease groupings with more than 15,000 first-time hospital admissions.

#### Acute hospital admission

Health records were also linked to 30,054,191 acute hospital admissions, of which 17,264,379 were dated between 2001 and 2016 and could be matched (Table 1—Source Data 1 & 2).

### Disease classification

The main diagnosis of acute hospital admission records was used to classify records into disease categories, which corresponded to disease blocks as described in the chapters of the ICD10(12). In order to model the effect of disease incidence, we used only the first admission of disease for each individual, excluding subsequent visits to the hospital for the same disease. The term “incidence” is used throughout this study to refer to the first recorded hospital admission of the disease during the study period.

### Design

Mortality trends were modelled using morbidity trends: we first determined the major disease categories (ICD10 blocks) associated with the most lives lost by taking into account the frequency of the disease (as measured by hospitalisation) and its effect on survival (as measured by the subsequent all-cause mortality of patients admitted for the disease compared to the mortality of everyone else). We combined these two measures to calculate a burden of death weighting for each block. We then looked at how the age-adjusted trends in hospitalisation rates (as a proxy for incidence) changed for each disease, by decade of birth, projecting that if incidence of a disease fell by a given percentage, its contribution to mortality would fall similarly. We then calculated a weighted average change in hospitalisation rates, reflecting the expected effect of all measured disease incidence changes on mortality rates, driven by decade of birth. Similarly, we looked at how the (age-adjusted) 5-year survival rates following first hospitalisation changed by year of hospitalisation. For each block this again gives a contribution towards reduced mortality, and the weighted average, the expected effect of changes in survival of the combined diseases on overall mortality. Adding these effects (and noting we assessed changes in survival from incidences over one decade), gives the expected effect on overall mortality from decade of birth to subsequent decade of birth from the effect of changes in disease incidence and survival, under the (necessarily simplified) model that incidence is a function of birth cohort and survival post incidence is a function of year of incidence.

### Statistical analysis

#### Mortality

A Cox proportional hazards model using NRS mortality data – fitting sex, decade of birth, and deprivation – was used to quantify mortality in the Scottish population during the study period. The same analysis was run stratified by sex and deprivation. Unless otherwise stated, (for example median age differences in Kaplan-Meier curves), years of life of a hazard effect have been calculated by multiplying the lnHR by 10 (ref. 13). Only individuals with complete records were included in the analysis.

#### Morbidity

We grouped the main diagnoses of each NSS hospital admission into categories, as laid out by the ICD10 Chapters, and included only the first instance of admission for a category per individual (discarding subsequent repeat visits to hospital for the same disease). Analysis was restricted to more common disease blocks. Visual inspection suggested a pragmatic threshold of at least 15,000 first-time admissions (see Table 2—Source Data 2 for all disease categories meeting this threshold).

Effects on the incidence of hospitalisation for the more common disease blocks was quantified using Cox proportional hazard models based on age, with events defined as the first incidence of hospitalisation. We fitted sex, deprivation, and decade of birth as covariates. Again, the same analyses were performed stratifying by sex and deprivation.

In order to quantify all-cause mortality in the five years following hospitalisation, person-time of individuals was divided into phases, corresponding to the study start until hospitalisation, the first five years after hospitalisation, and the remaining time in the study. For example, an individual admitted to hospital in 2004 for ischaemic heart disease I20-25 (IHD) and surviving until 2010 would contribute three phases to the model: one for the period until hospitalisation (no event), one for the first 5 years after hospitalisation (no event), and one >5 years after hospitalisation (event after 1 year). The status of each phase was fitted as a covariate in a Cox proportional hazards model(14) with death as the event, adjusting for sex and deprivation:

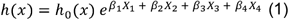

Where h_0_ is the baseline hazard, x the patient age, and X_1_-X_4_ the covariates sex, deprivation, and logically coded phase status (0-5 years True/False and >5 years True/False), with corresponding effect sizes *β*_1_-*β*_4_. This yielded estimates of the proportional hazard of status (0-5 and >5 years) after hospitalisation compared to pre-hospitalisation mortality. The same model was run, stratified by sex and deprivation.

#### Burden

For disease blocks with at least 15,000 first admissions during the study period, the relative mortality burden of each disease block was calculated as the excess mortality (lnHR) in the years after hospital admission (Equation 1) multiplied by the number of first-time admissions for the disease block, as follows:

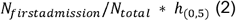

Where N_firstadmission_ is the total number of first hospital admissions of the disease during the study period, N_total_ is the total number of individuals in the study, and h_(0,5)_ is the mortality of individuals in the first five years following hospitalisation compared to individuals who were never hospitalised for the disease, measured in lnHR. The resulting value was then scaled to [0-1] and provides a relative measure of the number of lives lost due to the disease, with higher values indicating a disease which is more common or associated with higher subsequent mortality, and lower values indicating a disease which is rare or associated with lower subsequent mortality. Whilst this measure may in principle be affected by differing age patterns on incidence, it was judged sufficient for our purpose – to establish broad relative weightings of the importance of each disease.

To maintain a feasible computational burden within the national safe haven, subsequent analysis was restricted to the 25 blocks with the highest burden of death on the population (Table 2). We added C50-C50 malignant neoplasm of the breast, C60-C63, malignant neoplasms of male genital organs, and G30-G32 other diseases of the nervous system to this list, out of specific interest: in the sex-specific effects and awareness of the limitations of our method for Alzheimer’s disease (see discussion). All further analyses were performed on these top 28 blocks (T-28). The use of (first) hospitalisation for a disease as our definition of incidence is imperfect (e.g. for Alzheimer’s disease where hospitalisation following incidence is rare or delayed, and even first diagnosis in the community will often be preceded by a long latent period)(15).

#### Disease survival

Improvements in disease outcomes by ICD10 block were calculated by comparing 5-year all-cause mortality (Equation 1) following hospitalisation in 2001 with 5-year all-cause mortality following hospitalisation in 2011. We calculated 5-year mortality following first-time hospitalisation every year from 2001 to 2011 (i.e. mortality of patients admitted in that year) and fitted a 3^rd^ order polynomial regression to these estimates to calculate the underlying change in mortality following hospitalisation between 2001 and 2011.

#### Mortality estimates from morbidity

Estimates of the improvement in incidence of hospitalisation between decades of birth was combined into an overall improvement by performing a weighted sum of all diseases, with weights derived from the relative burden of death of each disease (see above).

### Ethical approval

This study was approved by the Public Benefit and Privacy Panel for Health and Social Care under application number 1617-0255/Joshi. As clinical records are provided without explicit patient consent, the panel requires the public benefit of the research to clearly outweigh any impact on individual patient privacy, and appropriate safeguards and security to be in place to protect patients. The panel granted access to de-identified patient data, accessible only through the National Safe Haven and only by University of Edinburgh researchers with data safeguarding qualifications. In addition, all results were reviewed by eDRIS before extraction from the safe haven to ensure no potentially identifiable information was made public.

### Patient and public involvement

Patients and the public were not involved in the study or its design, beyond their contribution of health records. Due to the retrospective study design and anonymised nature of the records, it was not feasible to contact individual patients nor involve them in the dissemination of results.

## Results

### Mortality

The population consisted of 1,967,130 Scottish individuals aged 35 years or older at the start of the study period (1 December 2000). 53.3% were female, 78.5% had been admitted to hospital at least once within the study period, and 30.6% died over the course of the study (31 January 2016). See Table 1 for detailed population characteristics.

Quantifying mortality effects using Cox proportional hazard models, we observed statistically significant associations (P < 1⨯10^−26^) between mortality and sex, deprivation and decade of birth (S Figure 2). Women showed lower overall age-adjusted mortality rates compared to men (70.7%; 95% CI 70.3%–71.1%), corresponding to an expectation of life of 3.5 years longer than their male counterparts, whilst individuals from the most deprived areas (top decile) suffered mortality rates more than twice as severe (207%; 95% CI 204%–209%) as those from the least deprived areas (bottom decile), corresponding to a difference in around 7 years of life. Median survival of men and women in the most deprived areas was 71.1 and 76.6 years, respectively, compared to 82.2 and 85.2 in the least deprived areas. A wide gap between the most deprived decile and the adjacent one for men is apparent visually: the difference in median survival between deprivation deciles 1 to 9 is roughly constant (0.82/1.05 years per decile for women/men), but moving from the 9th to 10th deprivation decile has a greater effect, especially for men (1.99/2.67 years for women/men). Lastly, individuals born in the decade commencing 1935 had age-adjusted mortality rates 245% (95% CI 239%–251%) of those born three decades later, corresponding to a difference in life expectancy of around 9 years of life.

### Morbidities and consequent mortality

Multiplying total number of hospitalisations during the study period (as a proxy for disease prevalence) by 5-year mortality after hospital admission (as a proxy for disease severity) provided a weight for the death burden of hospitalisation of each ICD10 block. We restricted our analyses to 28 of the top disease blocks for burden of death (T-28, see methods). Among the T-28, total cases of disease incidence (i.e. first-time admissions) during the study period ranged from 33,613 (A30-A49, Other bacterial disease) to 225,504 (R00-R09, Symptoms and signs involving the circulatory and respiratory systems) (Table 2). Per-person total cases of disease incidence (not age-adjusted) were 68.0% higher for the most deprived decile (188,905 individuals with 331,701 first-time admissions) compared to the least deprived decile (187,193 individuals with 195,617 first-time admissions). Between sexes, per-person incidence was 2.2% higher for men (917,788 individuals with 1,257,417 first-time admissions) compared to women (1,049,342 individuals with 1,407,223 first-time admissions) (Table 2—Source Data 1). In the first five years, all-cause mortality HRs ranged from 21 for patients admitted for C76-C80 (Malignant neoplasms of ill defined, secondary and unspecified sites; 23,490 cases) compared to all-cause mortality for those not admitted for C76-C80, to 1.8 for those admitted for K20-K31 (Diseases of oesophagus, stomach and duodenum; 128,000 cases) compared to all-cause mortality for those not admitted for K20-K31 during the study period.

Apart from sex-specific cancers, we observe significant differences in burden of death between men and women for injuries to the hip and thigh (S70-S79) and head (S00-S09), with the former having a higher burden in women due to more female cases and the latter having a higher burden in men due to more male cases. For both disease blocks, the effect of hospitalisation on subsequent mortality is greater in men than women (S70-S79 HR men: 3.19, women 2.44; S00-S09 HR men: 2.32, women 1.88 HR). Strikingly, 5-year mortality after hospital admission for IHD is higher for women (HR 2.01/1.70 women/men), but this is offset by the lower prevalence of hospitalisation in women (Table 2—Source Data 1).

### Trends in disease

To understand changes in disease survival rates, we next modelled the effects of a disease on all-cause mortality by year of hospital admission for admissions between 2001 and 2011 and 5-year survival subsequent to admission. We find an overall improvement over time in patient survival following hospitalisation, with a median decline between 2001 and 2011 in the 5-year HR of 16.8% for admitted cases across the T-28 diseases. The biggest improvements were for malignant neoplasms of the breast (C50) and male genital organs (C60-C63), which have seen 68.7% (95% CI 64.1%–72.7%) and 50.2% (95% CI 42.9%– 56.6%) declines in the 5-year hazard ratio between 2001 and 2011 mortality, respectively. On the other hand, other acute lower respiratory infections (J20-J22) and other diseases of the urinary system (N30-N39) have seen increases in mortality hazard of 11.3% (95% CI 7.0%–15.7%) and 24.0% (95% CI 14.6%–34.1%), respectively (Table 2, S Figure 3).

We next modelled age-adjusted incidence of hospitalisation for a disease by birth decade, under the simplified model that incidence is a cohort rather than period effect – essentially modelling that current incidence is the effect of (previous) lifetime exposures, rather than current exposures. We find disease incidence has fallen decade on decade of birth for cancers, cardiovascular, and intestinal diseases, but this improvement appears to have slowed down in the last decade of birth (1955-1965) considered. Age-adjusted incidence of influenza and pneumonia (J09-J18) and other bacterial diseases (A30-A49) has worsened by decade on decade of birth, over the whole range of births considered (S Figure 4).

When taking both trends in incidence and survival into account – adding 1) the average age-adjusted incidence rate reductions between decade on decade of birth to 2) the 2001-2011 reductions in 5-year disease mortality – we observe the death burden of cancers is declining most (Figure 1a). Notably, breast and prostate cancers have seen the largest improvement of all disease categories in the last decade. Other diseases of the urinary system (N30-N39), other bacterial diseases (A30-A49), other acute lower respiratory infections (J20-J22), and influenza and pneumonia (J09-J18) have all seen increases in their effect on age-adjusted all-cause mortality.

Overall, we see broad consistency in the scale of improvements across decades of birth, except for malignant neoplasms of respiratory and thoracic organs (C30-C39), where we see greater decade-on-decade improvements amongst later decades (Figure 1b). Averaging these individual disease effects on death, using burden of death weightings, we can then compare the modelled death rates with those observed, and see broad correspondence, with the 1935 and 1945 decades, showing the greatest improvements. Overall, our morbidity model suggests an average decade of birth on decade of birth lnHR of death of 0.30 (∼3 years of life) (Table 2).

The shape of disease modelled mortality improvements by decade of birth broadly tracks the observed changes. Furthermore, when stratified by sex a similar effect is seen: apparent distinctions in projected morbidity driven mortality are associated with distinctions with observed mortality (correlation 0.89); i.e. mortality trends in the study can largely be explained by trends in disease incidence and survival (S Table 1; S Figure 6). Across sex and deprivation strata, taking into account disease survival improvements between 2001 and 2011 and all improvements in disease incidence between decades of birth, we find the largest reductions in death are due to improvements in ischaemic heart diseases (I20-I25), malignant neoplasms of digestive organs (C15-C26), and malignant neoplasm of respiratory and intrathoracic organs (C30-C39), while the largest increases in death are due to other bacterial diseases (A30-A49) and influenza and pneumonia (J09-J18). In addition, the deterioration in other diseases of the urinary system (N30-N39) morbidity shows a consistent increase with deprivation, while other diseases of the digestive system (K90-K93) shows consistently larger improvements in more deprived classes (Table 2—Source Data 1).

Overall, we estimate 61.2% (95% CI 29.9%–92.6%) of the improvement in mortality rates was due to improvements in survival following hospital admission, with the balance arising from reduced (age-adjusted) admission rates (Table 2). Improved outcomes for cancers were particularly driven by post-admission survival, especially C60-C63 (88% of mortality improvement attributable to survival rather than incidence), C50 (85%) and C76-80 (68%), whereas for cardiovascular diseases, the balance was more even I80-89 (52%), I60-I69 (46%), I30-I32 (43%), I20-I25 (38%).

As previously noted, disease severity was defined as the log hazard ratio for subsequent all-cause mortality among those with a previous admission for an index group of conditions compared with those with no such admission. We regarded the rate of improvement in disease severity over time as being constant if there was the same relative fall in log hazard rate over successive time periods (so for example we regarded a fall in lnHR from 0.6 to 0.3 as equivalent to a fall from 0.3 to 0.15). Assuming the improvements in survival following hospitalisation continue for the coming decade, and differences between incidence in birth cohorts remains the same, we project a 21% slowing of improvements in mortality (−0.242 lnHR c.f. −0.305 lnHR; Table 3). Essentially, at least arithmetically, the population mortality benefits from improved cancer treatments in 2001-2011 will be hard to repeat as so much benefit has already accrued. Admittedly, this is a consequence of our model: essentially judging it equally difficult to reduce 50 excess deaths following cancer hospital admission associated to 25, as it was to reduce from 100 to 50, and as such should be considered speculative. On the other hand - our model is clearly valid *in extremis*: if all excess cancer deaths were eliminated, no further cancer driven improvement in mortality would be possible.

**Table 3:** Mean (over birth decades) decade of birth on decade of birth improvements in morbidity for the study period, and projections into the subsequent decade by sex and deprivation. Mortality improvements were estimated from morbidity records by combining the mean improvement in hospitalisation rate across birth cohorts and the improvement in disease severity between 2001 and 2011. This was then projected forward assuming improvements in age-adjusted hospitalisation rate between birth cohorts remained constant and improvements in severity remained proportional to the (now reduced) overall mortality of the disease group.

| Stratified | Group | Current Improvements |  |  | Projected Improvements |  |  |
| --- | --- | --- | --- | --- | --- | --- | --- |
|  |  | Hospital Admission Rate (lnHR) | Five Year Mortality After Admission (lnHR) | Combined (lnHR) | Hospital Admission Rate (lnHR) | Five Year Mortality After Admission (lnHR) | Combined (lnHR) |
| None |  | -0.1182 | -0.1866 | -0.3047 | -0.1182 | -0.1235 | -0.2418 |
| Sex | M | -0.1428 | -0.1913 | -0.3340 | -0.1428 | -0.1273 | -0.2701 |
| Sex | F | -0.0971 | -0.1823 | -0.2794 | -0.0971 | -0.1130 | -0.2101 |
| SIMD | 1 | -0.1154 | -0.2204 | -0.3360 | -0.1154 | -0.1280 | -0.2433 |
| SIMD | 2 | -0.1157 | -0.1592 | -0.2743 | -0.1157 | -0.0456 | -0.1610 |
| SIMD | 3 | -0.1202 | -0.1186 | -0.2392 | -0.1202 | -0.0370 | -0.1572 |
| SIMD | 4 | -0.1418 | -0.1759 | -0.3201 | -0.1418 | -0.0541 | -0.1982 |
| SIMD | 5 | -0.1231 | -0.1731 | -0.2958 | -0.1231 | -0.0914 | -0.2145 |
| SIMD | 6 | -0.1305 | -0.1928 | -0.3227 | -0.1305 | -0.0998 | -0.2303 |
| SIMD | 7 | -0.1233 | -0.1787 | -0.3044 | -0.1233 | -0.1109 | -0.2343 |
| SIMD | 8 | -0.1305 | -0.2097 | -0.3402 | -0.1305 | -0.1422 | -0.2726 |
| SIMD | 9 | -0.0926 | -0.1619 | -0.2546 | -0.0926 | -0.1033 | -0.1959 |
| SIMD | 10 | -0.0950 | -0.1970 | -0.2920 | -0.0950 | -0.1092 | -0.2043 |

## Discussion

In a study of 1,967,120 lives and 10,718,084 hospital admissions, we observed a median age at death of 82.2/85.2 for men/women in the highest socioeconomic decile, and 11.1/8.6 years less for the lowest decile. Cancers (C), cardiovascular disease (I), respiratory diseases (J), and unclassified symptoms and signs (R) were the principal ICD10 chapters recurring in the top 28 disease blocks where hospital admission was associated with the greatest subsequent all-cause mortality, which was a product of the rate of first hospital admission with group of conditions and of all-cause mortality in the five years following admission with that condition. Specifically, our top five causes of hospitalisations associated with subsequent burden of all-cause deaths were, in descending order, ‘Influenza and pneumonia’ (more common and with higher subsequent mortality than the average T-28 disease), ‘Symptoms and signs involving the circulatory and respiratory systems’ (common), ‘Malignant neoplasm of respiratory and intrathoracic organs’ (higher mortality),’ Symptoms and signs involving the digestive system and abdomen’ (common), and ‘General symptoms and signs’ (common and higher mortality). Whilst the latter might appear a benign diagnosis, our results suggest it is a fairly strong and frequent marker of subsequent all-cause mortality.

Across decades of birth, we modelled a reduction in mortality hazard of 0.737 (95% CI 0.730–0.745) due to improvements in morbidity, which broadly tracked improvements in observed mortality. The modelled improvement was 61% accounted for by reduction in excess mortality subsequent to admission and 39% accounted for by a fall in incidences of disease (as measured by hospital admission rates). The important (i.e. burden-of-death weighted) improvements in incidence were driven by cancers and heart disease, whilst improvement in outcomes following admission were mostly driven by cancer, particularly breast and prostate cancer. In contrast, we found deteriorations in the incidence of bacterial disease and in mortality following admission for respiratory and urinary infections. Levels of morbidity and mortality varied strongly across socioeconomic groups, but patterns in **changes** of such were generally less apparent. Men showed greater rates of improvement in mortality and morbidity than women, with lung and throat cancers contributing most to male improvements and IHD contributing most to female improvements.

In conclusion, we find trends in morbidity appear to partly explain trends in mortality. The progress in prevention and cure within oncology and prevention of heart disease account for the greatest parts of mortality improvement in 2001-2016, and our model suggests mortality improvements may slow, simply because the absolute effect of progress in treatment of these diseases will be difficult to repeat. However, there is scope for further improvements in life expectancy, especially if new progress is made in the treatment of other diseases associated with death, or if prevention initiatives accelerate.

### Strengths and weaknesses of the study

This study has avoided some of the known issues with cause of death recording(16) since it does not use cause-specific mortality and tracks wider disease effects and subsequent mortality (such as frailty) beyond direct causes of death, by combining hospitalisation and death records. Implicit tracking of underlying causes through an associated effect (admission to hospital for a disease) may improve estimates of trends in mortality, even if the underlying cause is obscure. We are also able to partition trends in deaths due to a disease based on trends in prevalence and incidence, which has been done for IHD(17), but not simultaneously across diseases in the same dataset. Also, our results are unaffected by population shifts as we excluded immigrants into Scotland after 2001, and instead reflect trends within the defined groups. Combined with the scale of our data, this consistent tracking has enabled us to make like-for-like comparisons of the mortality outcomes of different disease classes across socioeconomic groups and their trends over time.

However, this study also has a number of limitations. Whilst necessary given the data structure, excluding all migrants during the study period will in many cases have excluded healthy people moving for economic or lifestyle opportunity. When health status influences the propensity to migrate, it forms a competing risk for both mortality and morbidity, which can bias results(18). Nevertheless, as we are principally concerned in the trend of morbidity and mortality, only changes in migration biases – not their absolute level – should affect our results materially.

Secondly, we used recorded main position coding of first hospital admission of a disease block as the index event. Hospital admission is only a proxy for the incidence of severe disease, and likely works better for chronic conditions like breast cancer where there is a single disease process that can result in a number of hospital admissions and may result in mortality in the timeframe of the study. It works perhaps less well for acute conditions, such as influenza, which can have distinct occurrences over the study period, and it fails to capture chronic conditions which are managed in general practice or the community but eventually lead to death, such as dementia and multiple sclerosis. Also, association between hospital admission for a condition and the latter occurrence of death does not necessarily imply causation, and may be confounded by other health risks, such as socioeconomic status, a second disease, or behavioural factors, although our analysis stratified by socioeconomic status may partly mitigate these effects. Modelling comorbidities simultaneously or modelling the causal effects of one morbidity on another and consequently on death has been beyond the scope of this study. In addition, our model assumed that (age-adjusted) incidence is a function of year of birth, but that mortality post-incidence is a function of the year of incidence. This is clearly an oversimplification, but necessary given that year of birth and year of incidence are completely confounded for a given age at incidence.

By relying on routine data, we have been able to create a very large dataset, at the expense of being unable to externally check individual fields, such as primary diagnosis, where there are known to be inaccuracies(19). However, our conclusions will be less affected if these inaccuracies are reasonably stable over time. Furthermore, we have used a simple definition of burden of disease: the total number of cases multiplied by the excess hazard across all ages. More complex definitions might have looked at Years of Life Lost (YLL) and taken into account any age-related pattern in incidence. However, our measure is simple and is predominantly used to weight the diseases and especially their improvements to calculate an overall improvement in all-disease impact, rather than as an absolute measure of burden.

Lastly, our model assumed disease incidence and survival hazards were proportionate. Incidence of hospital admission and death used “age” as the baseline hazard determinant, while mortality following admission used “time since admission”. In practice we do not expect the risks analysed to be truly proportionate, nonetheless we consider the analysis of trends to be revealing. Conversely, recently doubt was cast on the effect of using decade of birth groupings in a standardised mortality analysis(20) due to changing patterns of births within the decade: that concern does not apply here as the Cox baseline hazard used full (not rounded) age information, with decade of birth fitted as a covariate.

Changes in hospital admission policy have likely affected changes in observed hospitalisation rate. Conversely, survival rates could be influenced if for example, only more serious cases were admitted. One example from our study is the observed increase in incidence of influenza and pneumonia, which was offset by increased survival and may therefore reflect changes in coding practices and/or frequency of referral(21). Some degree of caution is needed in observing the changes in morbidity, although such biases will have tended to offset each other in terms of the overall change in the mortality arising from morbidity for such a disease block. However, clearly the partition of effect between incidence and survival can be affected.

### Strengths and weaknesses in relation to other studies

There was a degree of correspondence in the principal burdens assessed here and a recent study by the Scottish Burden of Disease study (SBD)(22). This study used the same population and the same study period but assessed YLL (weighting young deaths more as opposed to our method which counted all deaths equally), included individuals younger than35 years old, and used different disease groupings. Their principal burdens were IHD (ranked 13^th^ in our list of burdens), tracheal, bronchus and lung cancers (3^rd^), chronic obstructive pulmonary disease (12^th^), stroke (10^th^), and Alzheimer’s disease (–). Aside from Alzheimer’s disease, discussed below, much of the distinction appears to arise from our observation of an association between death and admissions with indistinct diagnosis (not considered a valid specific cause of death by SBD). In the case of influenza and pneumonia, differences arise due to our study identifying a marker of frailty as well as a direct cause of death, combined with SBD grouping influenza and pneumonia under lower respiratory infections. A relative strength of our study stems from usage of incident morbidity (as marked by hospitalisation) in advance of death, based on recorded diagnosis at the time of hospital visit, thus tracking remote effects such as long term frailty rather than cause of death (which has known limited accuracy, particularly at older ages(16)). However, the principal strength arises from the ability to distinguish trends in incidence of morbidity from trends in subsequent survival. On the other hand, a relative weakness is that we are reliant on hospital admission as a marker of incidence; therefore, diagnosed or latent (presumably milder) cases in the absence of admission are not visible to us, leading for example to significant discrepancy with SBD in the apparent relative burden of Alzheimer’s disease, likely due to an understatement of its importance in our results.

The closing gap in mortality between the sexes and its widening across social classes observed in our study is consistent with recent findings from the Office of National Statistics, summarised by Torjesen(23), which looked at socioeconomic deprivation in England and Wales. Similarly, a recent study of health inequality in England found rising levels of lifespan inequality across socioeconomic groupings arising from increasing inequalities across a broad span of causes of death(24). These studies had the advantage of a larger sample size (∼7.5 million deaths cf. 600,000 in our study) and could therefore track trends in mortality and cause of death between stratified groups more accurately. However, Scotland’s unique linkage of death records and electronic health records through eDRIS allowed us to directly model changes disease mortality at an individual level (avoiding issues with cause of death recordings and shifts in population demographics). Our study has the advantage of partly explaining these trends in mortality inequality through changes in disease incidence and survival: men experienced greater improvements in incidence of lung cancer and survival following heart disease hospitalisation compared to women, while more socially deprived individuals (men and women combined) suffered worse deteriorations in infectious disease, especially for the incidence and survival of hospitalisation for urinary tract infections. However, in contrast to Bennet *et al*.(24), we do not find a clear pattern in overall morbidity improvements across socioeconomic deciles in Scotland, and we do not observe a widening inequality in cancer, respiratory and Alzheimer’s disease morbidity within our study population, although we are underpowered to detect the latter and our disease groupings were not identical.

Lastly, a recent study of coronary heart disease mortality in Scotland, using a sophisticated model to apportion improvement between prevention and treatment, found improvements for coronary heart disease between 2001-2010 were similar across social classes, and reported 33%–61% of these improvements could be attributed to advances in treatment(17). Given the very different methods, albeit studying the same population, there is reasonable concordance with our own study: we find roughly equal improvements in heart disease across social classes and estimate 38% (95% CI 28%–48%) of these improvements stem from increased survival after hospitalisation for ischaemic heart disease. Hotchkiss *et al*.(17) are able to further partition improvements by uptake of primary and secondary prevention drugs and treatments. Such detailed analysis of specific diseases has been beyond the scope of our study.

### Implications for clinicians and policymakers

Much of the improvements in mortality observed in Scotland between 2001–2016 can be attributed to reductions in morbidity, as captured by hospital admissions. While this study examined mortality and morbidity in the Scottish population only, there is a substantial concordance in mortality trends across high-income countries(7), as well as similarities in disease-related mortality trends between Scotland and the rest of the UK(6), warranting similar studies to be performed in other high-income countries. It is a testament to healthcare services that the majority of mortality improvements appear to stem from advances in disease survival post-admission. Observed improvements in cancer incidence and survival – especially breast and prostate – coincide with a continued effort within Scotland(25), the UK(26), and other high-income nations(27) to improve prevention and care of these diseases. However, the rapid advances in survival of both heart disease and cancer modelled by our study between 2001 and 2011 will be hard to continue to the same extent, as so much progress has already been made. At the same time, the observed deteriorations in infectious disease coincide with global increases in antimicrobial resistance(28) and emphasise the need to prioritise research in this area: infectious disease will become a larger contributor to mortality and may contribute to a widening of health inequalities between socioeconomic classes. If these current trends in morbidity continue, we expect morbidity-driven improvements in mortality to slow down. However, the life expectancy gap between Scotland and other high-income countries(29) suggests further mortality improvements are possible. The rate of this improvement will hinge upon whether advances in all major diseases categories – especially infectious disease – can catch up with the progress we have recently seen on heart disease and cancer, and whether preventable deaths from external causes (such as suicide and drug-related deaths), which cannot be accurately tracked using hospital admissions, decrease rather than rise.

## Data Availability

Individual de-identified participant data cannot be shared. Statistical code and technical details are available upon request from the corresponding author.

## Funding

The study was funded by the Lloyds Banking Group, for the creation, curation and dissemination of knowledge in the public interest, in particular to improve estimates of future population size and morbidity and mortality rates to facilitate healthcare and other government planning. All analyses stratified by socioeconomic deprivation (Table 2, Table 3, Supplementary Table 1, and Supplementary Data Files 1-5) were confidentially re-analysed using 10 socioeconomic groups specified by Lloyds, rather than the Scottish Index of Multiple Deprivation. No Lloyds employees were granted access to individual patient data. Data access was granted to University of Edinburgh researchers only and only through the national safe haven by the Public Benefit and Privacy Panel for Health and Social Care under application number 1617-0255/Joshi.

The study and its design were conceived by the last author. The funder reviewed the design and said that adding stratification by socioeconomic status was a key requirement for meaningful analysis and their funding was conditional upon this, a request to which the authors readily agreed. The funder was kept informed of interim analyses and reviewed the draft manuscript, occasionally suggesting additional analyses or requesting clarifications. The funders were not permitted to and did not request the removal of any results.

Authors affiliated with eDRIS and NHS Scotland were compensated by PKJ using the grant from Lloyds Banking Group, in line with normal pricing for the work undertaken. PRHJT was funded by the MRC Doctoral Training Programme (MR/N013166/1) and the University of Edinburgh College of Medicine and Veterinary Medicine. JFW was funded by the MRC QTL in health and disease. Apart from Lloyds Banking Group, the funders had no role in the study design, data collection, analysis, and interpretation, or the decision to submit the work for publication.

## Acknowledgements

We would like to thank Lloyds for the funding and in particular Craig Butler and Stuart McDonald for their engagement with the project. We would also like to thank Steve Pavis and Doug Kidd for their guidance with the Public Benefit and Privacy Panel for Health and Social Care.

## Competing interests

PKJ reports grants from Lloyds Banking Group, PLC, during the conduct of the study. This grant was in part used to pay for research costs of JJK and CMF. The remaining authors declare no competing interests.

## List of Tables and Figures

**Table 1—Source Data 1: Study descriptives by sex, deprivation, and deceased status (Excel Document)**. SIMD – Scottish Index of Multiple Deprivation; Age Entry – Age at the start of the study period (1 December 2000); Age Exit – Age at the end of study period (31 January 2016) or at the end of life; Admissions – Records of hospital admission, where Count represents the number of individuals with at least one hospital admission record, Mean/Median/SD represent the mean/median/standard deviation of number of hospital admission records per individual, and Sum represents the total number of admission records during the study period across all individuals; A-Z – First letter of the ICD10 main condition recorded at all hospital visits; SD – Standard Deviation

**Table 1—Source Data 2: Count of records within each ICD10 block (Plain Text Document)**. Main diagnoses of hospital admission records were grouped into ICD 10 chapter. For each ICD 10 chapter, counts of the 3-letter ICD 10 subcategories with 10 or more admissions are reported.

**Table 2—Source Data 1: Disease mortality burden by sex and deprivation (Excel Document)**. Contains sheets for 1) total incident cases, 2) mortality in the first 5 years following disease admission, 3) mortality after 5 years following disease admission, 4) the ratio of 0-5 and >5 year mortalities, 5) the relative burden of disease, and 6) the weight of each disease, by sex and deprivation. SIMD – Decile of Scottish Index of Multiple Deprivation, from least deprived (1) to most deprived (10).

**Table 2—Source Data 2: Relative mortality burden of all disease groupings with at least 15**,**000 first-time admissions (Excel Document)**. ICD10 – Diseases contained within the disease grouping, coded by International Classification of Disease Codes, Tenth Revision. See (Table 1—Source Data 2 Disease Count) for counts of 3-letter ICD10 records within each ICD10 block. Disease – Description of the diseases within the disease grouping. Hospital visits – Number of first-time admissions with main diagnosis falling within the disease block. Mortality – Mortality compared to individuals who had not yet or ever been admitted for the disease group. Relative Burden of Death – Ratio of hospital admissions to total individuals, multiplied by the mortality in the first five years after admission. Weight burden of death rescaled to a total value of 1.

## Supplementary Material

**S Figure 1:**
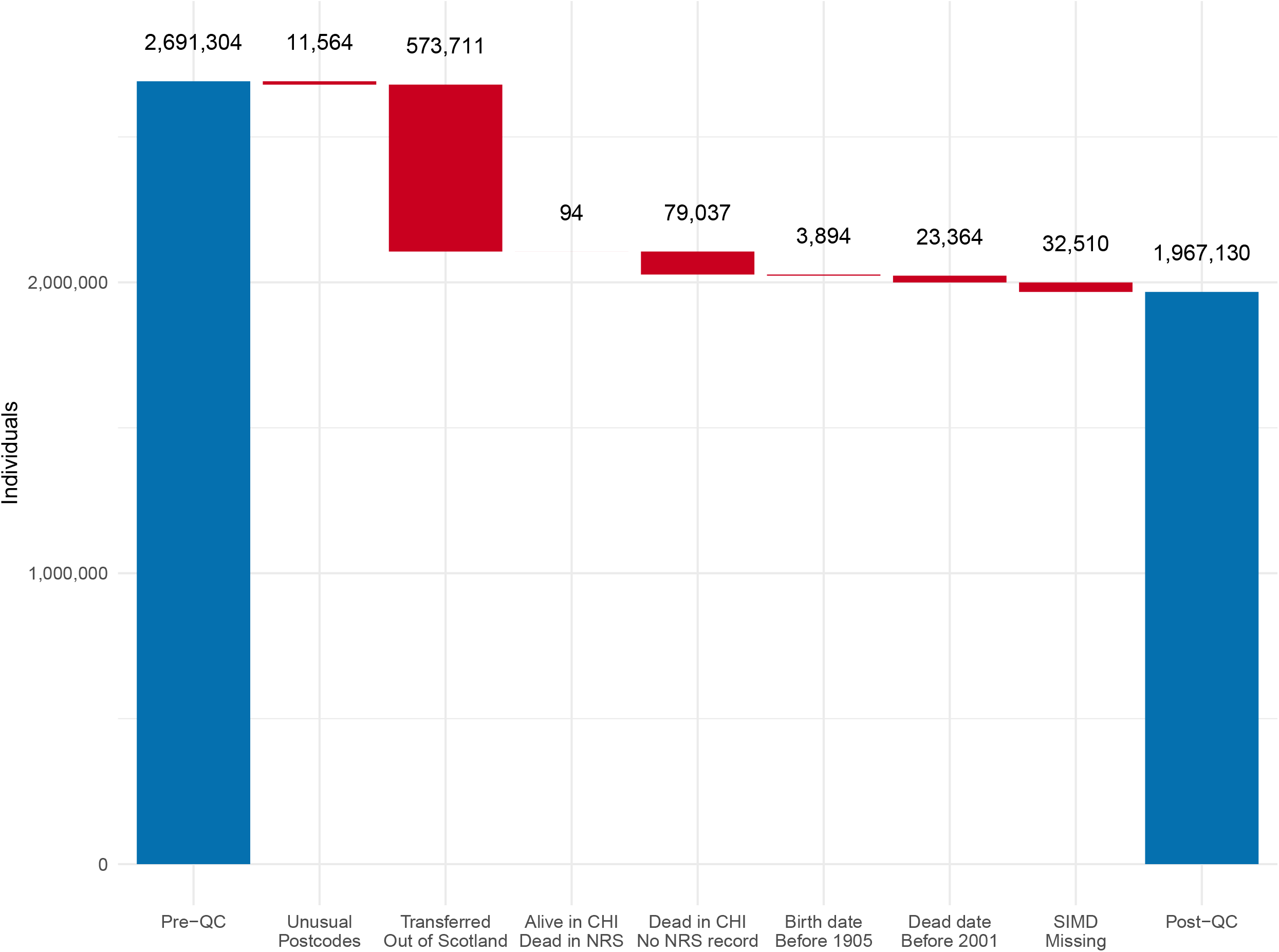
Waterfall chart of individuals excluded from the study during quality control. We started with ∼2.7 million primary care records from the NSS Community Health Index (CHI). Records were excluded from individuals with uncommon district postcodes (N < 5000), individuals missing from the CHI database in 2016 (and therefore likely transferred out of Scotland), individuals with record discrepancies between the CHI and National Records of Scotland databases, and individuals with records outside of the study dates or missing information on socioeconomic class.

**S Figure 1—Source Data 1: Details of quality control performed on the dataset (Excel Document)**. Primary care records from the National Health Services Scotland underwent multiple rounds of quality control. For each step, descriptives of the excluded individuals are provided. Unusual postcode – District-level postcodes shared between fewer than 5,000 individuals (anomalous postcodes, often with special meanings, such as “marketing campaign”). Transferred out of Scotland – Individuals absent from NSS records at the end of the study period, suggesting transfer out of Scotland or otherwise poor tracking. CHI – Community Health Index. NRS – National Records of Scotland.

**S Figure 2:**
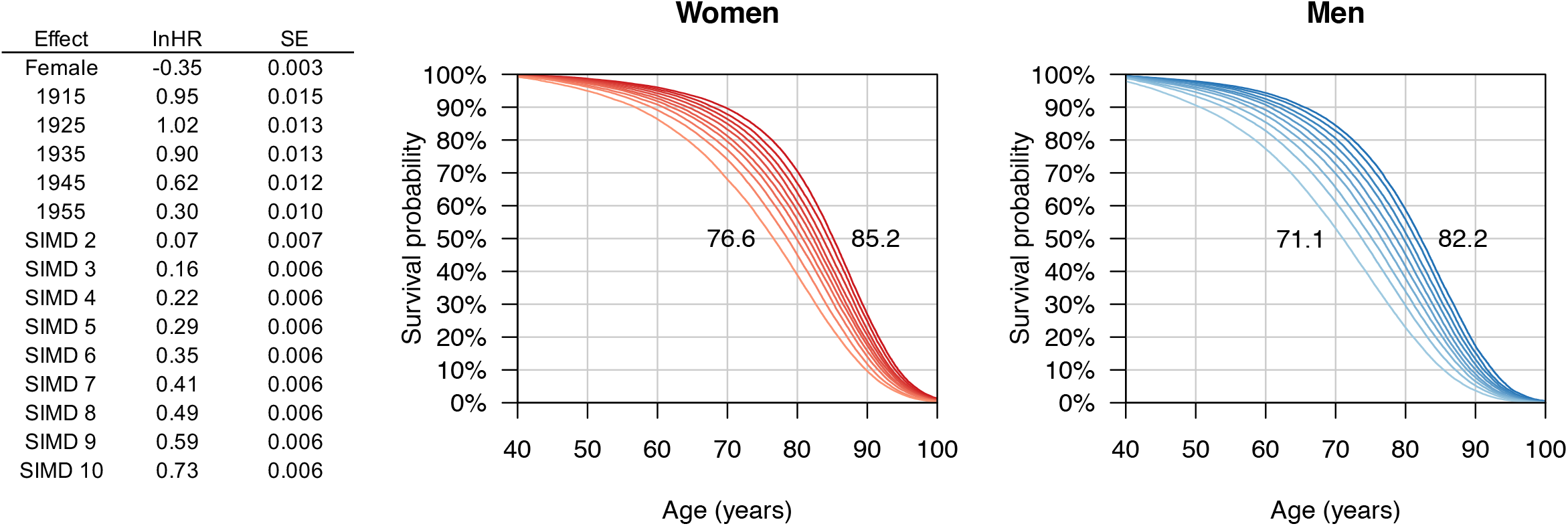
Kaplan-Meier survival curves show ∼10 year differences in median survival between top and bottom deprivation deciles. Empirical survival curves were calculated from 1,967,130 individuals and 647,752 mortality records between 2001–2016, with median survival annotated for the least and most deprived deciles. The accompanying table quantifies the relative mortality of the sexes, birth cohorts, and deprivation deciles in log hazard ratios, with SIMD 1 males from the 1965 birth cohort as reference. See S Figure 2—Source Data 1 for numerical details of the Kaplan-Meier curves including median survival of each decile.

**S Figure 2—Source Data 1: Kaplan-Meier survival curves show ∼10 year differences in median survival between top and bottom deprivation deciles (Excel Document)**. SIMD – Scottish Index for Multiple Deprivation decile; Records – Total number of individuals in the decile; N Max – Total number of uncensored individuals during the study period; Events – Number of individuals who died during the study period; Mean – Mean survival time; SE – Standard error of the mean survival time; Median – median survival time (i.e. number of years 50% of individuals survive); 95% CI – 95% confidence interval around the median survival time (years).

**S Figure 3:**
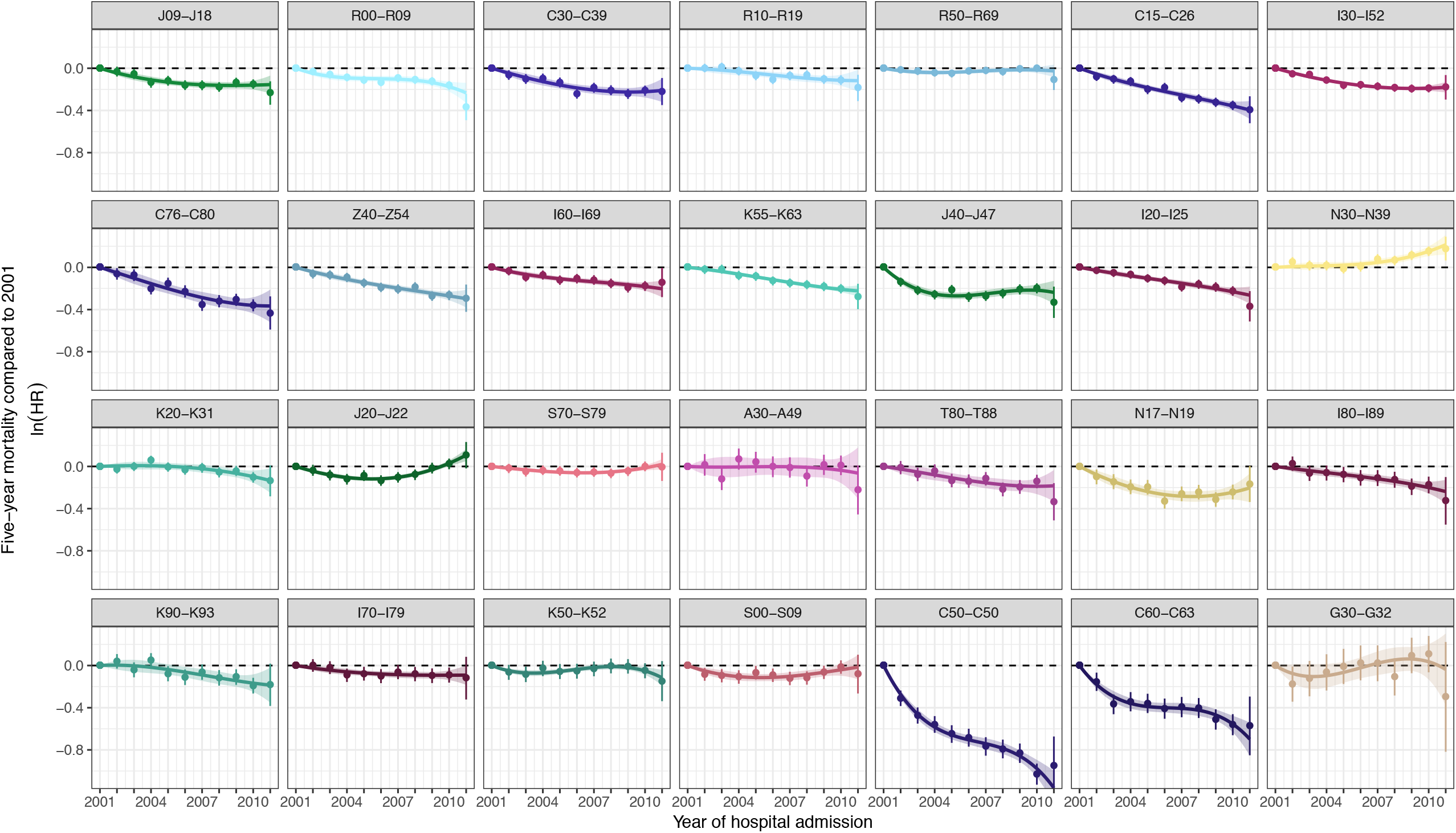
Age-adjusted reductions in excess mortality following hospital admission between 2001–2011. Dots represent excess mortality in the first five years after hospital admission compared non-admissions, compared to the excess mortality in 2001, expressed in age-adjusted ln HR. The black dots represent five-year mortality following survival to the current year. Fitted are fixed intercept 3^rd^ power polynomials, weighted by inverse variance of estimates. The polynomial fitted value at 2011 with its standard error are used in morbidity driven mortality improvement models. See Supplementary Data File 3 for this graph stratified by sex and deprivation.

**S Figure 4:**
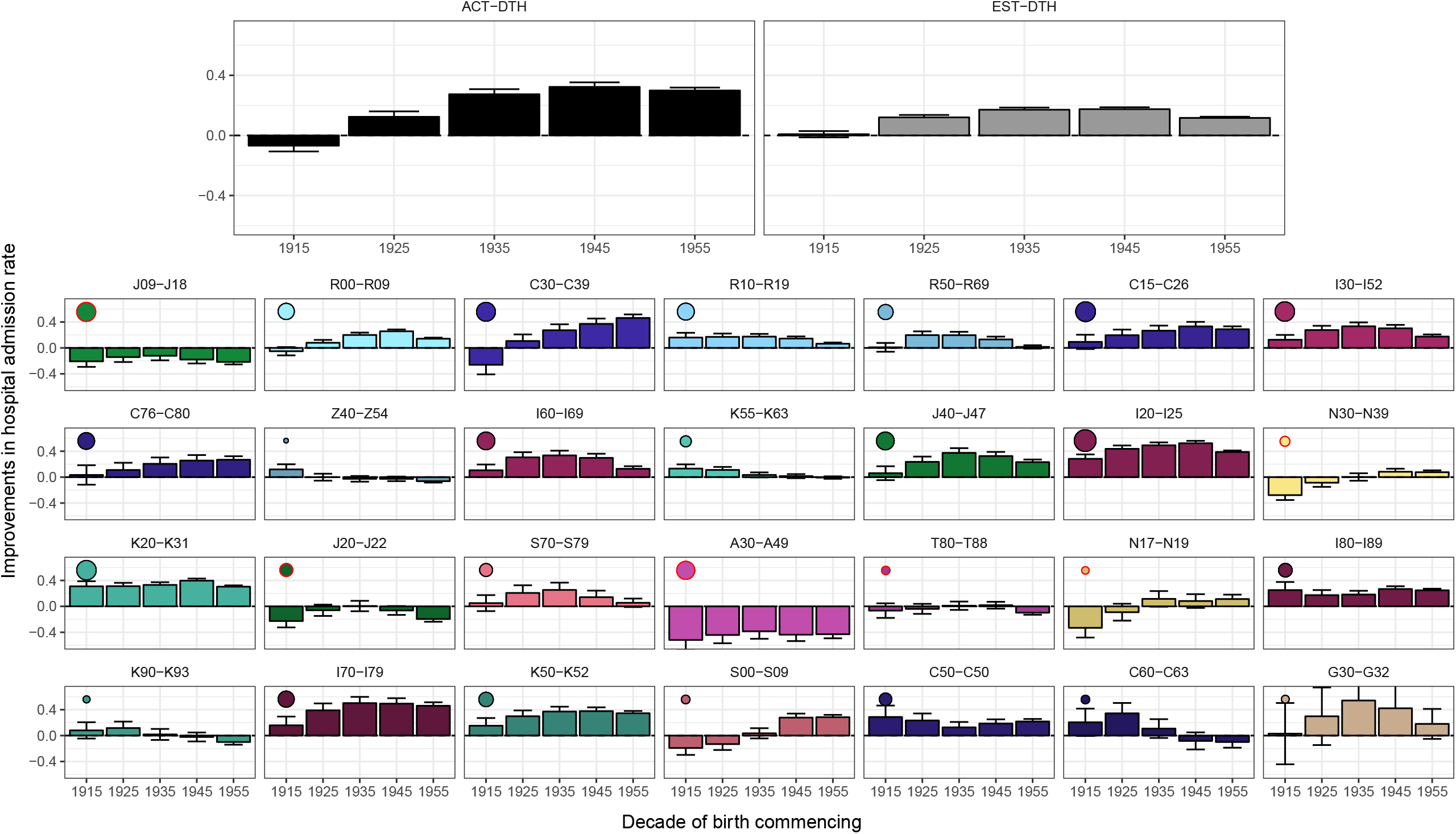
Age-adjusted reductions in hospital admission rate by decade of birth. Bars represent the improvements in hospital admission rate since the previous decade of birth, expressed in age-adjusted lnHR. Dots represent the relative contribution of the disease category to the overall (ie. thee burden of death weighted) improvements in mortality due to changing hospital admission rates. Larger dots indicate a greater contribution. A red circle around the dot indicates a negative contribution (i.e. deterioration). These overall improvements are shown in the grey bars - which represent the modelled effect of morbidity incidence on mortality. The black bars represent the improvements in mortality observed from death records, by decade of birth. See Supplementary Data File 4 for this graph stratified by sex and deprivation.

**S Figure 5:**
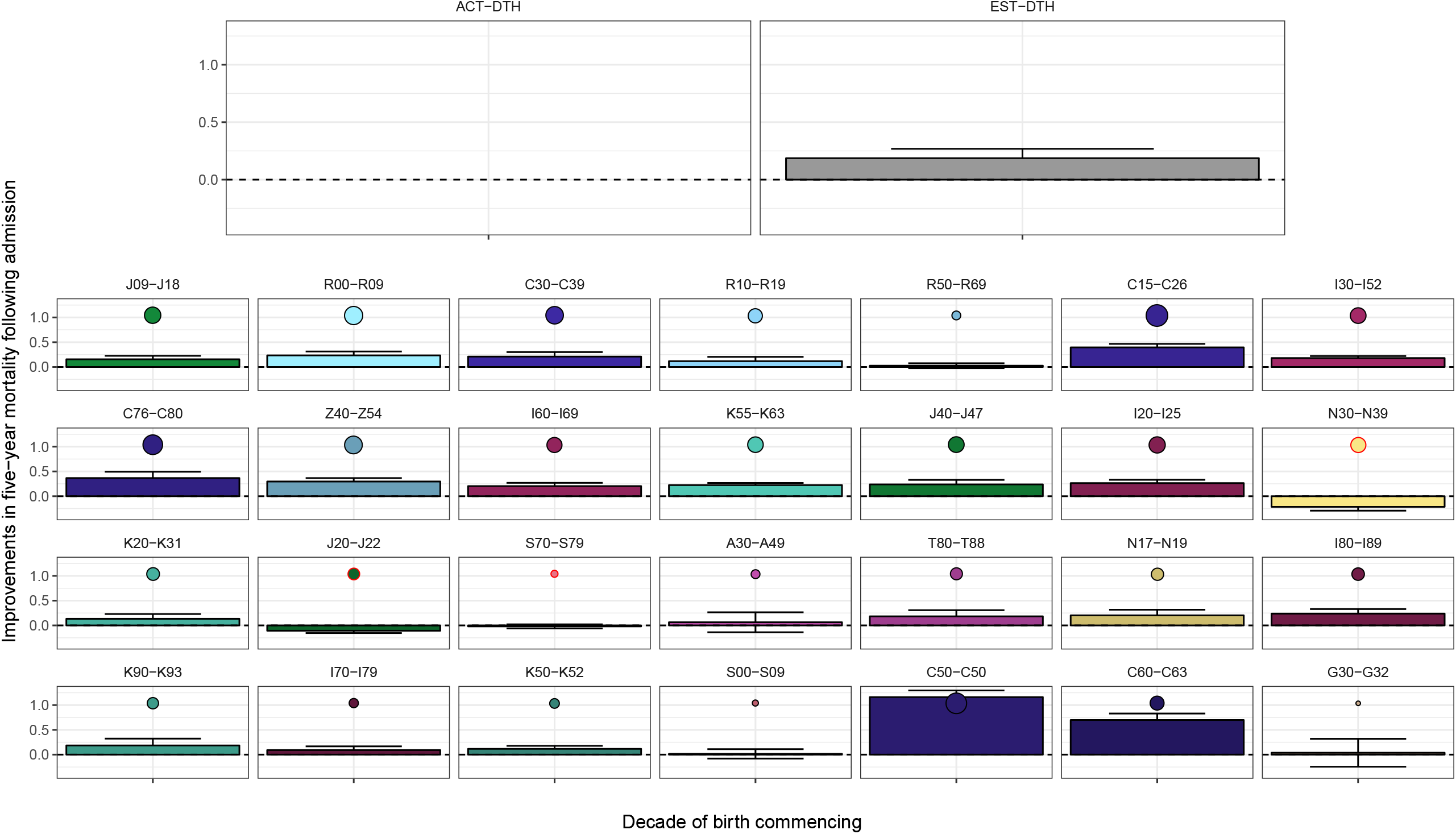
Age-adjusted improvements in survival following hospitalisation. Bars represent the age adjusted ln HR of survival for admissions in 2011 compared with 2001. Dots represent the relative contribution of the disease category to the overall (i.e. the burden of death weighted) improvements in mortality due to changing hospital admission rates. Larger dots indicate a greater contribution. A red circle around the dot indicates a negative contribution (i.e. deterioration). These overall improvements are shown in the grey bar - which represents the modelled effect of morbidity survival on mortality 2001 vs 2011. (As is obvious, the effect of death on death itself - first panel - is unchanged) See Supplementary Data File 5 for this graph stratified by sex and deprivation.

**S Figure 6:**
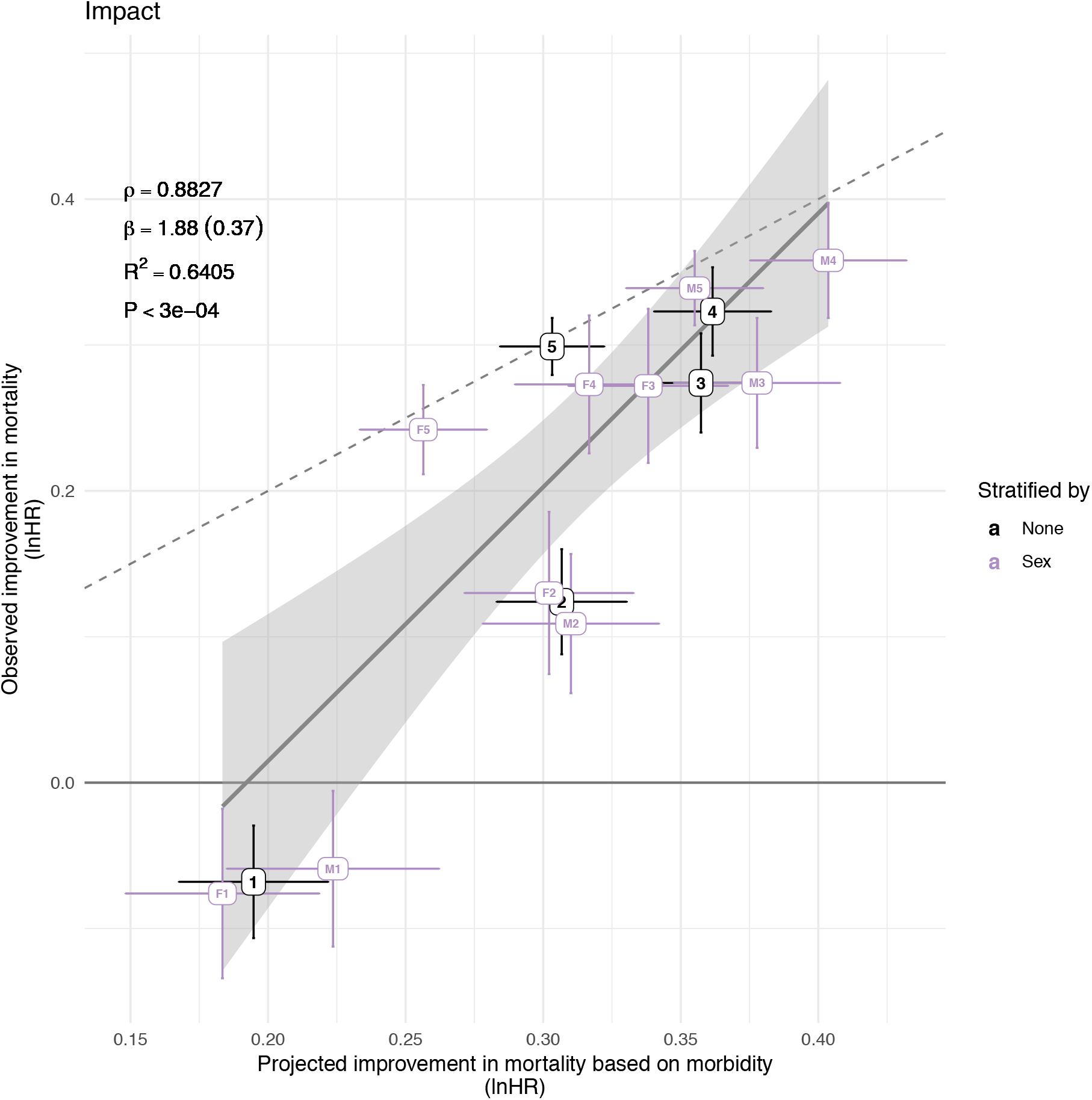
Improvements in mortality projected from changes in disease morbidity partially explain observed improvements in mortality. Improvements in morbidity, based on reductions in hospitalisation rate and 5-year mortality, were calculated for individuals stratified by sex and decade of birth. The dotted line represents perfect concordance between projected and observed mortality improvements, with points above this line indicating mortality improvements have been greater than projected from disease improvements, and points below this line indicating the opposite. The solid line is the regression of projected mortality on observed mortality and includes all shown data points, weighted by their uncertainty in both directions. Annotated are the regression slope (standard error), correlation, variance explained, and significance. Concordance between projected and observed mortality appears to increase with later decades of birth. M – Male; F – Female; 1 – improvements of the cohort born in the decade after 1915 compared with the cohort born in the decade before; 2 – 1925; 3 – 1935; 4 – 1945; 5 – 1955.

**S Table 1: Total Improvements in mortality and morbidity across all birth decades by sex and deprivation (Excel Document)**. Mortality improvements were estimated from morbidity records by combining the improvement in hospitalisation rate for the 1955–1965 cohort compared to the 1905– 1915 cohort and the improvement in disease severity between 2001 and 2011. Actual mortality improvements of the 1955–1965 cohort vs. 1905–1915 cohort was calculated directly from mortality records. SIMD – Scottish Index of Multiple Deprivation. Lives Saved – Disease groups which have benefited the most and least from the improvements in morbidity, by taking into account the burden of death and its improvement.

**Supplementary Data File 1 (Excel Document):** Improvements in incidence of hospitalisation and 5-year mortality following hospitalisation by sex and deprivation. Contains improvements in hospital admission rate, disease survival, and combined impact on mortality for each disease grouping, stratified by sex and deprivation.

**Supplementary Data File 2 (PDF document):** Age-adjusted reductions in combined hospital admission rate and survival following hospitalisation by decade of birth, stratified by sex and deprivation

**Supplementary Data File 3 (PDF document):** Age-adjusted reductions in excess mortality following hospital admission between 2001–2011, stratified by sex and deprivation.

**Supplementary Data File 4 (PDF Document):** Age-adjusted reductions in hospital admission rate by decade of birth, stratified by sex and deprivation.

**Supplementary Data File 5 (PDF Document):** Age-adjusted increases in survival following hospitalisation, stratified by sex and deprivation.

## Notes

### Author Declarations

All relevant ethical guidelines have been followed and any necessary IRB and/or ethics committee approvals have been obtained.

Any clinical trials involved have been registered with an ICMJE-approved registry such as ClinicalTrials.gov and the trial ID is included in the manuscript.

